# Varenicline as a Repurposable Drug Candidate for ADRD Prevention

**DOI:** 10.64898/2026.09.17.26363232

**Authors:** Phillip Ma, Jiaxu Zhou, Yan Cheng, John E. McGeary, Tracey H. Taveira, Ali Ahmed, Wen-Chih Wu, Edward Zamrini, Brittany N. Dugger, Louise Nicole C. Sevilla, Joel Anthony Nations, Yijun Shao, Ying Yin, Debby W. Tsuang, Mark W. Logue, Siamack Ayandeh, Charles Faselis, Stuart J. Nelson, Qing Zeng-Treitler

## Abstract

**Objective:** Effective strategies to prevent Alzheimer’s disease and related dementias (ADRD) are limited. Implementing our Medication-Wide Association Study (MWAS+), we highlighted varenicline as a candidate. This study evaluated the association between varenicline use and risk of ADRD relative to nicotine replacement therapy (NRT).

**Methods:** Using the All of Us Research Database, we identified adults aged 50 years or older with available lifestyle survey responses regarding tobacco use history who initiated smoking cessation therapy with either varenicline or NRT between 2006 and 2023. We limited our cohort to incident monotherapy users and designated the index date as the first prescription. Individuals with baseline ADRD or no follow-up were excluded. Baseline characteristics included demographics, comorbidities, BMI, and prior medication exposures. Outcomes were incident ADRD and a composite outcome of ADRD or death.

**Findings:** The cohort consisted of 9,584 individuals (1,801 varenicline users and 7,783 NRT users). After propensity score matching, there were 1,758 individuals in each group with balanced baseline characteristics. Varenicline use was associated with a reduced risk of ADRD (HR 0.516, 95% CI: 0.353–0.754) and the composite outcome ADRD or death (HR 0.657, 95% CI: 0.501–0.861). Kaplan–Meier curves also demonstrated varenicline’s sustained benefits through the study period.

**Implications:** Varenicline use is associated with a lower risk of ADRD among older adults receiving smoking cessation therapy. Further investigation of varenicline as a potential strategy for ADRD prevention is warranted. These findings underscored the value of explainable AI-guided approaches to drug repurposing.

**Highlight:**

- Artificial Intelligence (AI) Medication-Wide Association Study (MWAS+) identified varenicline as candidate for ADRD prevention.
- Varenicline linked to lower ADRD risk vs NRT in propensity score matched cohort.
- Consistent results for ADRD and composite outcome ADRD or death.

## 1. Introduction

Alzheimer’s disease and related dementias (ADRD) affect millions of individuals worldwide. The lack of broadly effective treatments for ADRD underscores the urgent need to focus on prevention strategies. This work centers on drug repurposing for ADRD prevention, leveraging a novel approach termed Medication-Wide Association Study Plus (MWAS+).^1^

The “MWAS” phase is hypothesis-free and mechanistically agnostic. We trained an AI model called hybrid value-aware transformer (HVAT)^2,3^ on approximately one million Veteran Affairs (VA) patient records, comprising 50% cases (patients who developed ADRD) and 50% controls (matched patients who did not develop ADRD in the given timeframe). The dataset captured up to 10 years of longitudinal clinical history, including medication prescriptions, comorbidities, laboratory results, procedures, and demographics.

The model was trained to distinguish cases from controls, after which an explainable AI method developed by our team^4^ was applied to identify drug exposures associated with reduced ADRD risk. This process generated several potential drug repurposing candidates. Among these, varenicline ranked second in its potential protective effect. A subsequent mechanistic review suggested that varenicline is a biologically plausible candidate, motivating a cohort study to evaluate its potential benefit in ADRD prevention.^5^

The cholinergic system is of strong interest for understanding Alzheimer’s pathophysiology and is a target for potential treatments. The loss of cholinergic neurons is a signature feature of Alzheimer’s disease that results in a decrease of acetylcholinergic neurotransmission, critical for memory and cognition.^6^ Cholinergic receptors include both muscarinic and nicotinic receptors.^7^ Smoking is a known risk factor for Alzheimer’s disease via numerous mechanisms including vascular damage, inflammation, and oxidative stress.^8^ Consequently, smoking cessation can reduce the risk of Alzheimer’s disease.^9^ Two FDA-approved treatments for smoking cessation are nicotine replacement therapy (NRT) and Chantix (varenicline).^10^ NRT may include patches, gum, and lozenges. All provide nicotine to reduce nicotine withdrawal and reduce the physiological impact of the 7000+ chemicals present in combusted tobacco.^11^ Varenicline is a partial agonist at α4β2 nicotinic receptors that was approved in 2006. Varenicline releases dopamine to reduce cravings and withdrawal while simultaneously blocking nicotine receptors.^12^ Varenicline has been shown to be more effective than NRT, and the combination of both shows synergistic positive effects on smoking cessation.^13^

In addition to being a partial agonist of the α4β2 nicotinic receptor, varenicline is a full agonist at the α7 receptor. Preclinical studies suggest that varenicline may have theoretical potential in Alzheimer’s Disease through multiple mechanisms, including nicotinic receptor modulation, synaptic plasticity enhancement, anti-inflammatory effects, and modulation of amyloid-beta (Aβ) activity.^14–17^ Despite these promising preclinical findings, clinical evidence remains limited, and the only Phase II trial has not supported these theoretical benefits. In a short-term, 6-week phase II crossover trial involving 66 patients with mild-to-moderate Alzheimer’s Disease, varenicline 1mg twice daily did not improve cognitive outcomes and was associated with an increased risk of adverse gastrointestinal effects.^18^ The lack of clinical benefit may reflect the short duration of treatment, small number of participants, and the inclusion of patients with established disease, a stage characterized by substantial neurodegeneration. Preclinical evidence suggests that nicotinic receptor modulation may be effective for prevention or for very early intervention rather than treatment of established disease.

In this plus (“+”) phase of our study, we are examining biologically plausible candidates through cohort studies. Because varenicline is used for smoking cessation, we employed a new-user design comparing varenicline with NRT as an active comparator. This study leveraged the NIH All of Us dataset,^19^ which contains rich baseline characteristics and longitudinal electronic health record data, including prescription information, enabling assessment of ADRD incidence associated with varenicline versus NRT.

## 2. Methods

### 2.1 Dataset

We conducted a cohort study using the All of Us Research database. We identified individuals with a history of tobacco use based on responses from the lifestyle survey (e.g., “Have you smoked at least 100 cigarettes in your entire life?”).^19^ Within this cohort, the three most commonly used smoking cessation medications were NRT (41.87%), bupropion (49.66%), and varenicline (8.47%). Since bupropion was also indicated for the treatment of depression and was not the focus of this study, we excluded individuals treated with bupropion. We further restricted the cohort to individuals who initiated either varenicline or NRT between 2006 and 2023. The year 2006 was selected as the start of the study period because varenicline was approved in that year. Additional inclusion criteria were age ≥50 years at treatment initiation, absence of ADRD at baseline, and at least one follow-up encounter after treatment initiation (**Figure 1**).

**Figure 1:**
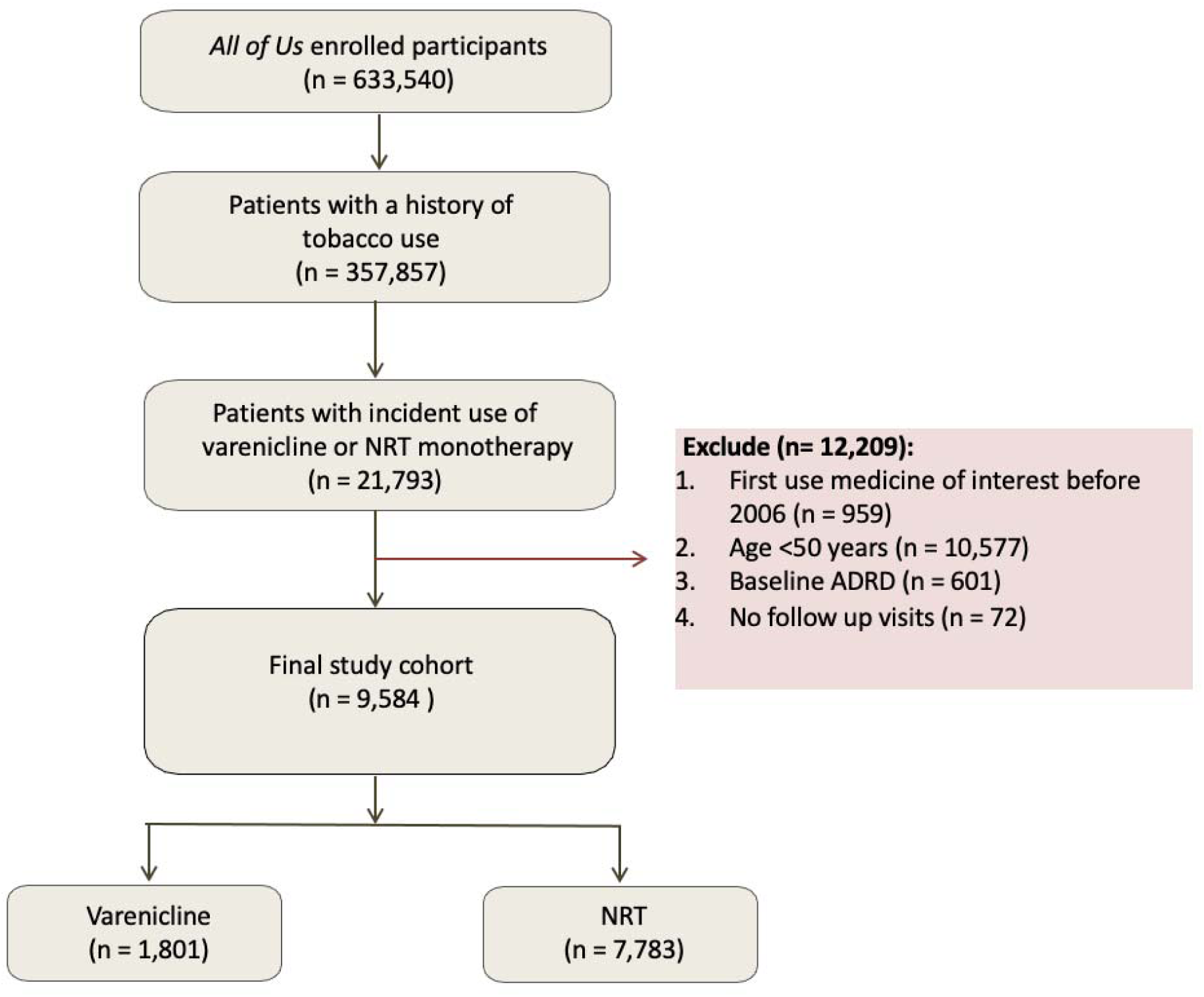
Cohort Assembly Flowchart.

### 2.2 Exposure

The primary exposure was initiation of varenicline versus NRT, identified from medication records. The index date was defined as the date of treatment initiation.

### 2.3 Outcomes

The primary outcome was incident ADRD, which was identified using International Classification of Diseases, Ninth and Tenth Revision (ICD-9 and ICD-10) diagnostic codes (**eTable 1**). ADRD was chosen as an endpoint as opposed to strict Alzheimer’s Disease due to the heterogenous use of ICD codes in this domain, which may not fully capture many instances of actual AD, as the only gold standard for AD diagnosis is after death with neuropathologic examination.^20,21^ In the primary analysis, death without a diagnosis of ADRD was treated as a censoring event. Since death may preclude the subsequent observation of ADRD, particularly among older adults, we additionally assessed a secondary composite outcome (ADRD or Death). Death served as an important competing risk for ADRD. This composite outcome addressed the potential for the individuals who died early to be at high risk for ADRD but not living long enough for diagnosis.

Participants were followed for up to 15 years from the index date until the first occurrence of ADRD, death, or last recorded encounter. We selected a 15-year follow-up horizon to capture long-term ADRD risk while minimizing bias from differential follow-up and sparse data at later time points.

### 2.4 Covariates

We included a set of baseline covariates measured at or before the index date, including comorbidities, demographics, vital signs, and prior medication exposures (**Table 1**).^22^ Chronic comorbid conditions were identified using ICD codes (**eTable 1**). The prior medication exposures were systematically selected based on their ingredients after excluding drug classes (e.g., antibiotics, hormones, electrolytes) that are not commonly associated with ADRD with a prioritization of psychoactive and other clinically relevant medications, especially those with properties or effects of anticholinergics. Finally, we retained the most frequently used medications.

**Table 1.**
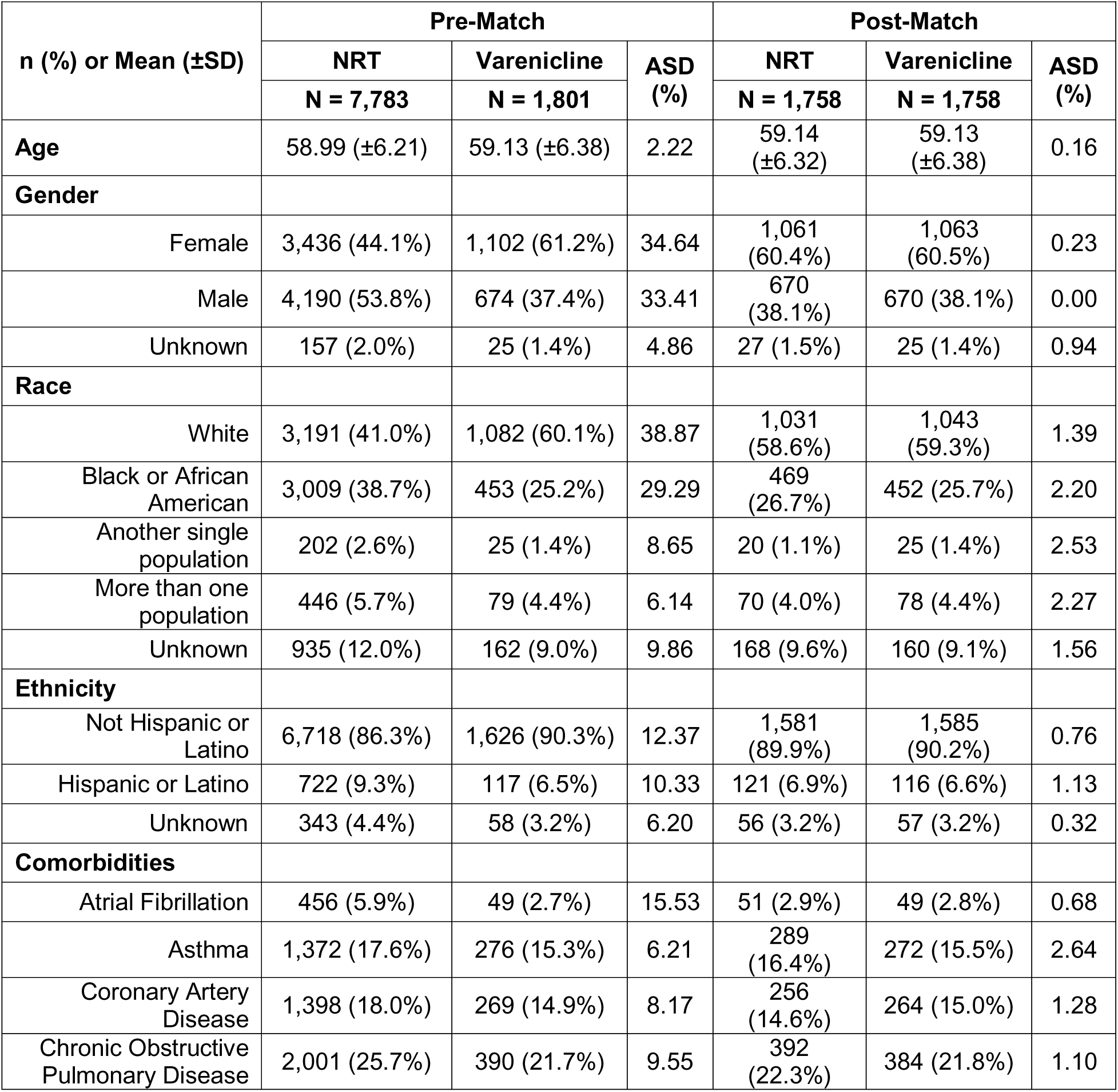
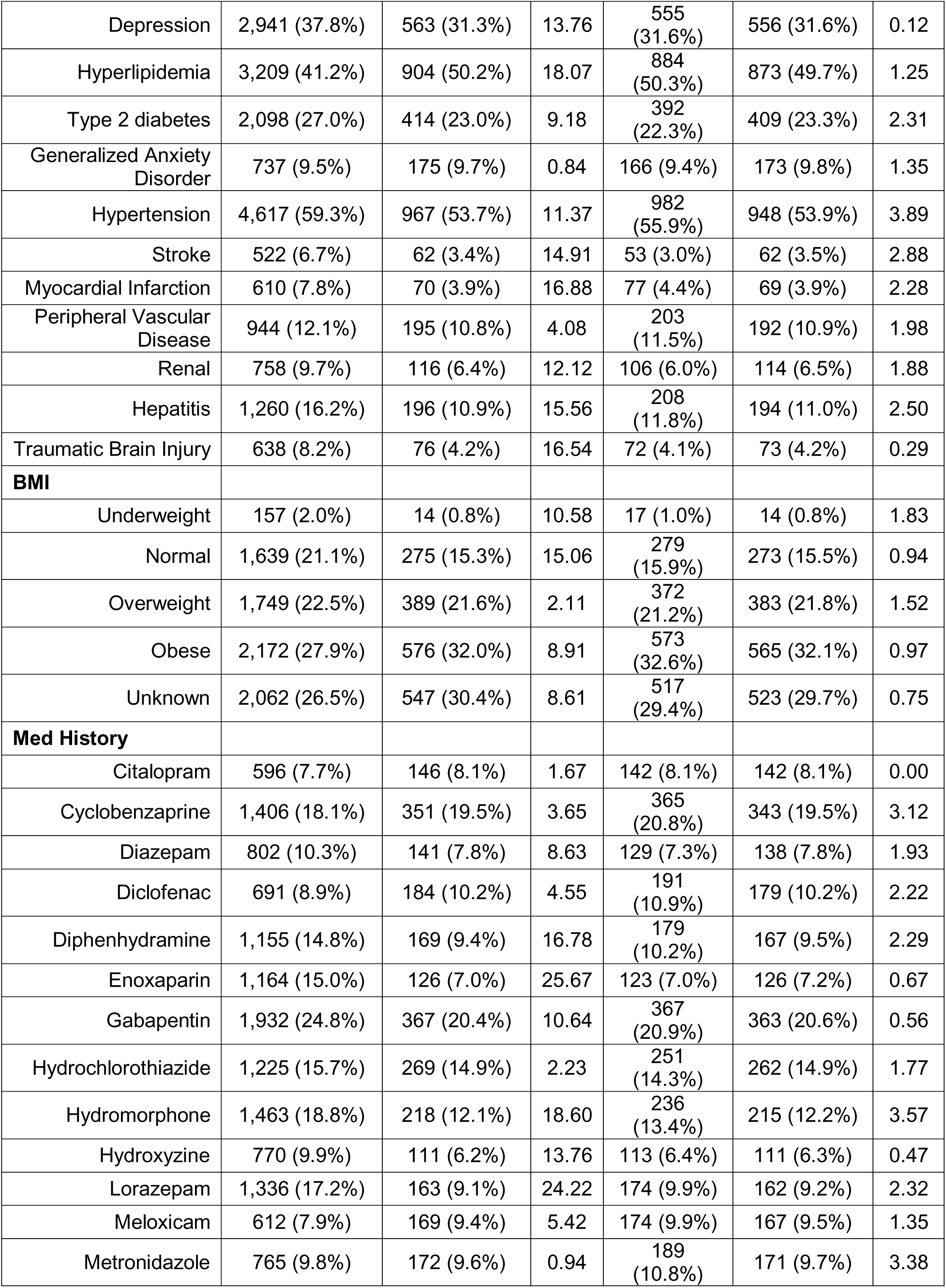
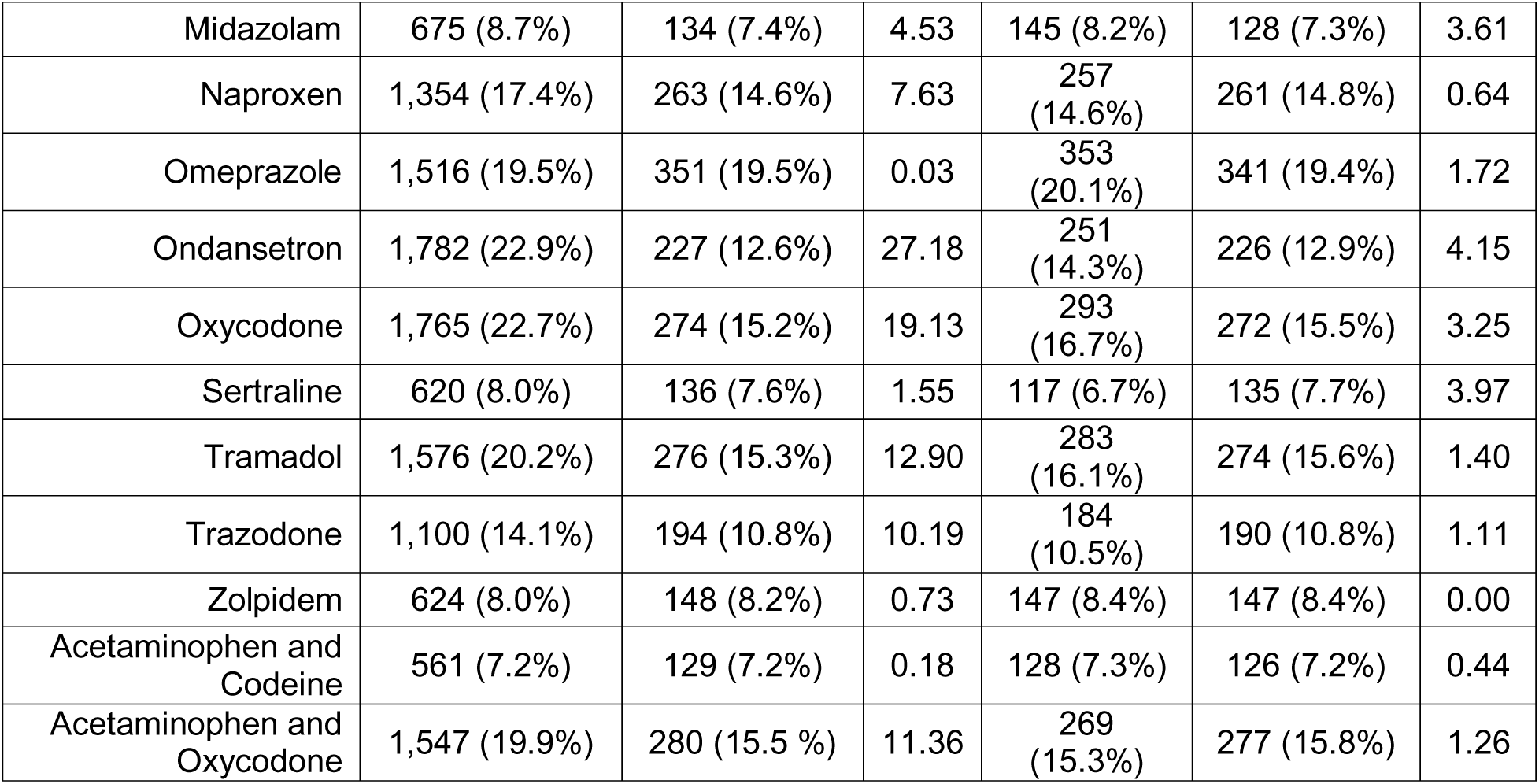
Baseline Characteristics: Varenicline vs. NRT.

### 2.5 Propensity Score Matching

We fit a logistic regression model to estimate the probability of initiating varenicline versus NRT using baseline covariates. Propensity score matching (PSM) was performed using the MatchIt package in R. Individuals in the varenicline and NRT groups were matched using nearest-neighbor matching with a caliper and without replacement. A caliper width of 0.2 standard deviations of the logit of the propensity score was used, consistent with commonly recommended practice to minimize residual bias while maintaining sufficient sample size.^23^

### 2.6 Statistical Analysis

#### 2.6.1 Descriptive Analysis

Baseline characteristics were summarized for both treatment groups before and after PSM. Covariate balance was assessed using absolute standardized differences (ASDs),^24^ calculated for both categorical and continuous variables, with an ASD <10% considered indicative of adequate balance. This approach was used to evaluate group comparability and the effectiveness of the matching procedure.

#### 2.6.2 Survival Analysis

We calculated hazard ratios (HRs) and 95% confidence intervals (CIs) for each outcome using Cox proportional hazards models implemented in the survival package (version 3.8.3) in R (version 4.5.0). Analyses were conducted in both propensity score matched and unmatched cohorts. In addition, Kaplan–Meier (KM) curves were generated to visualize time-to-event outcomes.

#### 2.6.3 Supplementary Analysis of Treatment Utilization and Post-Treatment Smoking Status

Treatment data after the index date were not included in the primary outcome analysis, consistent with the new-user design. To aid interpretation of our survival findings, we conducted supplementary descriptive analyses of treatment utilization patterns and post-treatment smoking status in the PSM cohort.

To assess treatment utilization, we calculated the number of prescriptions and cumulative drug exposure for each individual. Prescriptions were identified for both NRT and varenicline, regardless of specific product type (e.g. nicotine patch vs lozenge vs gum). Overlapping prescriptions and those separated by gaps of < 7 days were considered part of the same treatment episode, whereas a gap >7 days defined a new prescription episode. Cumulative exposure was defined as each individual’s total treatment duration.

We also assessed smoking status after treatment based on ICD-coded smoking-related diagnoses. Smoking status was defined as “Yes” if any smoking-related ICD diagnosis was recorded after treatment initiation and “No” otherwise.

## 3. Results

### 3.1 Cohort Assembly

A total of 633,540 participants enrolled in the All of Us Research Program were initially screened. Among them, 357,857 individuals had a documented history of tobacco use based on lifestyle survey responses. We identified 44,252 participants who used varenicline, NRT, or bupropion as smoking cessation medications and restricted the cohort to monotherapy incident users. Among these, 21,793 participants received prescriptions for either varenicline or NRT between 2006 and 2023. After applying study eligibility criteria, we excluded 12,209 individuals, including those with medication initiation before 2006 (n = 959), age younger than 50 years at treatment initiation (n = 10,577), baseline ADRD (n = 601), and no follow-up visits after treatment initiation (n = 72). The final analytic cohort consisted of 9,584 participants, including 1,801 varenicline users and 7,783 NRT users (**Figure 1**).

### 3.2 Baseline characteristics (before matching)

Our study cohort contained 7,783 individuals receiving NRT and 1,801 individuals receiving varenicline. Prior to matching, baseline characteristics differed between groups, with several variables showing meaningful imbalance ([ASD] > 10%) (**Table 1**).

Age was comparable between groups (mean: 58.99 vs. 59.13 years; ASD = 2.22%). However, the distribution of sex differed considerably, with higher proportion of females in the varenicline group (61.2% vs. 44.1%; ASD = 34.64%) and a lower proportion of males (37.4% vs. 53.8%; ASD = 33.41%). There was also variation in racial composition, with a greater proportion of White individuals in the varenicline group (60.1% vs. 41.0%; ASD = 38.87%) and a higher proportion of Black or African American individuals in the NRT group (38.7% vs. 25.2%; ASD = 29.29%). Differences in ethnicity were more modest but still apparent (not Hispanic or Latino: 90.3% vs. 86.3%; ASD = 12.37%).

Several comorbidities occurred more frequently in the NRT group, including atrial fibrillation, depression, hypertension, stroke, myocardial infarction, renal disease, hepatitis, and traumatic brain injury (all ASDs > 10%), with the exception of hyperlipidemia, which was more frequent in the varenicline group (ASD = 18.07%). Body mass index categories also showed disparities, particularly for the normal-weight category (21.1% vs. 15.3%; ASD = 15.06%).

Prior medication exposures differed notably between groups. Several medications, including diphenhydramine (ASD = 16.78%), enoxaparin (ASD = 25.67%), gabapentin (ASD = 10.64%), hydromorphone (ASD = 18.60%), hydroxyzine (ASD = 13.76%), lorazepam (ASD = 24.22%), ondansetron (ASD = 27.18%), oxycodone (ASD = 19.13%), tramadol (ASD = 12.90%), trazodone (ASD = 10.19%), and acetaminophen and oxycodone (ASD = 11.36%), were more commonly used in the NRT group.

### 3.3 Baseline Characteristics (Post-matching)

After PSM, there were 1,758 individuals in each group in our analytic sample, and baseline characteristics were well balanced (**Table 1**). All ASDs were < 10%, indicating good matching. Age and sex were comparable between groups (59.14 vs. 59.13 years; ASD = 0.16%, female: 60.4% vs. 60.5%; ASD = 0.23%). Racial and ethnic distributions were similar as well, with small differences across categories (all ASDs < 3%). Comorbidities, BMI, and prior medication exposures also achieved acceptable balance after matching (ASD <5%, <2%, and <5% across categories, respectively).

### 3.4 Survival Analysis (outcome of ADRD)

We used the Cox proportional hazards regression to explore the association between smoking cessation treatment and incident ADRD (**Table 2**). In the unmatched cohort, varenicline use was associated with a lower hazard of ADRD compared with NRT (HR = 0.575, 95% CI: 0.410– 0.806, p = 0.001), corresponding to a 42.5% reduction in hazard. After PSM, the association remained statistically significant and slightly stronger (HR = 0.516, 95% CI: 0.353–0.754, p = 0.001), corresponding to a 48.4% lower hazard of ADRD among varenicline users.

**Table 2.** Cox Regression Results for ADRD.

|  |  | Before PSM (ADRD) |  | After PSM (ADRD) |  |
| --- | --- | --- | --- | --- | --- |
|  |  | Adjusted HR<br>(95% CI) | Pr(>\|z\|) | HR<br>(95% CI) | Pr(>\|z\|) |
| **Therapy** | Varenicline vs NRT | 0.575 (0.410,0.806) | 0.001 | 0.516 (0.353,0.754) | 0.001 |
| **Age** | Age at index | 1.069 (1.050,1.088) | 0.000 |  |  |
| **Gender** |  |  |  |  |  |
|  | Female vs Male | 0.778 (0.613,0.987) | 0.039 |  |  |
|  | Unknown vs Male | 0.814 (0.359,1.849) | 0.623 |  |  |
| **Race** |  |  |  |  |  |
|  | Black or African American vs White | 1.383 (1.062,1.800) | 0.016 |  |  |
|  | Another single population vs White | 1.057 (0.508,2.199) | 0.882 |  |  |
|  | More than one population vs White | 0.785 (0.430,1.433) | 0.430 |  |  |
|  | Unknown vs White | 0.654 (0.287,1.488) | 0.312 |  |  |
| **Ethnicity** |  |  |  |  |  |
|  | Hispanic or Latino vs Not Hispanic or Latino | 2.322 (1.117,4.826) | 0.024 |  |  |
|  | Unknown vs Not Hispanic or Latino | 1.721 (0.650,4.557) | 0.274 |  |  |
| **BMI** |  |  |  |  |  |
|  | Unknown vs Normal | 0.774 (0.568,1.055) | 0.105 |  |  |
|  | Obese vs Normal | 0.591 (0.421,0.829) | 0.002 |  |  |
|  | Overweight vs Normal | 0.814 (0.594,1.116) | 0.201 |  |  |
|  | Underweight vs Normal | 0.830 (0.335,2.059) | 0.688 |  |  |
| **Comorbidities** |  |  |  |  |  |
|  | Atrial Fibrillation | 1.170 (0.748,1.828) | 0.491 |  |  |
|  | Asthma | 1.051 (0.765,1.442) | 0.761 |  |  |
|  | Coronary Artery Disease | 1.245 (0.906,1.710) | 0.176 |  |  |
|  | Chronic Obstructive Pulmonary Disease | 1.183 (0.912,1.533) | 0.205 |  |  |
|  | Depression | 1.101 (0.846,1.433) | 0.475 |  |  |
|  | Generalized Anxiety Disorder | 0.820 (0.532,1.266) | 0.371 |  |  |
|  | Hepatitis | 0.978 (0.722,1.326) | 0.887 |  |  |
|  | Hypertension | 1.113 (0.851,1.457) | 0.435 |  |  |
|  | Hyperlipidemia | 0.871 (0.673,1.127) | 0.294 |  |  |
|  | Myocardial Infarction | 1.136 (0.759,1.700) | 0.536 |  |  |
|  | Peripheral Vascular Disease | 1.084 (0.781,1.503) | 0.630 |  |  |
|  | Renal | 0.919 (0.622,1.359) | 0.672 |  |  |
|  | Stroke | 1.755 (1.202,2.562) | 0.004 |  |  |
|  | Traumatic Brain Injury | 0.973 (0.639,1.481) | 0.899 |  |  |
|  | Type 2 Diabetes | 1.174 (0.898,1.534) | 0.240 |  |  |
| **Med History** |  |  |  |  |  |
|  | Gabapentin | 1.038 (0.771,1.398) | 0.805 |  |  |
|  | Acetaminophen and Codeine | 0.432 (0.253,0.739) | 0.002 |  |  |
|  | Acetaminophen and Oxycodone | 1.210 (0.897,1.633) | 0.212 |  |  |
|  | Citalopram | 1.347 (0.903,2.010) | 0.145 |  |  |
|  | Cyclobenzaprine | 0.962 (0.691,1.340) | 0.819 |  |  |
|  | Diazepam | 1.235 (0.853,1.790) | 0.264 |  |  |
|  | Diclofenac | 1.140 (0.738,1.762) | 0.555 |  |  |
|  | Diphenhydramine | 1.098 (0.774,1.558) | 0.600 |  |  |
|  | Enoxaparin | 0.720 (0.492,1.053) | 0.091 |  |  |
|  | Hydrochlorothiazide | 0.951 (0.697,1.298) | 0.751 |  |  |
|  | Hydromorphone | 0.836 (0.583,1.198) | 0.330 |  |  |
|  | Hydroxyzine | 1.362 (0.897,2.069) | 0.147 |  |  |
|  | Lorazepam | 1.077 (0.783,1.481) | 0.649 |  |  |
|  | Meloxicam | 0.354 (0.172,0.727) | 0.005 |  |  |
|  | Metronidazole | 0.743 (0.477,1.156) | 0.188 |  |  |
|  | Midazolam | 0.822 (0.526,1.285) | 0.390 |  |  |
|  | Naproxen | 0.685 (0.480,0.979) | 0.038 |  |  |
|  | Omeprazole | 1.058 (0.792,1.413) | 0.704 |  |  |
|  | Ondansetron | 1.116 (0.809,1.540) | 0.504 |  |  |
|  | Oxycodone | 1.230 (0.888,1.703) | 0.214 |  |  |
|  | Sertraline | 0.866 (0.543,1.381) | 0.545 |  |  |
|  | Tramadol | 1.096 (0.804,1.495) | 0.561 |  |  |
|  | Trazodone | 1.024 (0.716,1.465) | 0.897 |  |  |
|  | Zolpidem | 0.877 (0.579,1.328) | 0.535 |  |  |

Several covariates included in the pre-matching model also showed associations consistent with prior literature on ADRD risk. Higher age was strongly associated with higher hazard (HR = 1.069, 95% CI: 1.050–1.088, p < 0.001), consistent with the well-established relationship between advancing age and dementia.^22^ Female sex was associated with lower hazard (HR = 0.778, 95% CI: 0.613–0.987, p = 0.039). Black or African American individuals had a higher hazard compared with White individuals (HR = 1.383, 95% CI:1.062–1.800, p = 0.016), and Hispanic or Latino ethnicity was associated with higher hazard (HR = 2.322, 95% CI: 1.117– 4.826, p = 0.024).^22^

BMI showed an inverse association, with obesity associated with a lower hazard of ADRD (HR = 0.591, 95% CI: 0.421–0.829, p = 0.002).

Among the comorbidities, stroke was strongly associated with an increased risk of ADRD (HR = 1.755, 95% CI: 1.202–2.562, p = 0.004), whereas other conditions showed weak or non-significant associations.

Prior medication exposures showed mixed associations. Acetaminophen and codeine, meloxicam, and naproxen were associated with a lower risk of ADRD, while other medications were not significantly associated with the outcome.

### 3.5 Survival Analysis (composite outcome of ADRD and death)

A Cox proportional hazards regression model was also used to explore the association between smoking cessation treatment and the composite outcome of ADRD or death (**Table 3**).

**Table 3.** Cox Regression Results for Composite Outcomes.

|  |  | Before PSM<br>(ADRD or Death) |  | After PSM<br>(ADRD or Death) |  |
| --- | --- | --- | --- | --- | --- |
|  |  | Adjusted HR<br>(95% CI) | Pr(>\|z\|) | HR<br>(95% CI) | Pr(>\|z\|) |
| **Therapy** | Varenicline vs NRT | 0.660<br>(0.524,0.832) | 0.000 | 0.657<br>(0.501,0.861) | 0.002 |
| **Age** | Age at index | 1.061<br>(1.048,1.075) | 0.000 |  |  |
| **Gender** |  |  |  |  |  |
|  | Female vs Male | 0.825<br>(0.698,0.976) | 0.025 |  |  |
|  | Unknown vs Male | 0.858<br>(0.491,1.500) | 0.592 |  |  |
| **Race** |  |  |  |  |  |
|  | Black or African American vs White | 1.511<br>(1.254,1.820) | 0.000 |  |  |
|  | Another single population vs White | 1.106<br>(0.650,1.880) | 0.711 |  |  |
|  | More than one population vs White | 1.383<br>(0.979,1.954) | 0.066 |  |  |
|  | Unknown vs White | 0.768<br>(0.412,1.431) | 0.405 |  |  |
| **Ethnicity** |  |  |  |  |  |
|  | Hispanic or Latino vs Not Hispanic or Latino | 1.864<br>(1.077,3.227) | 0.026 |  |  |
|  | Unknown vs Not Hispanic or Latino | 1.517<br>(0.734,3.133) | 0.260 |  |  |
| **BMI** |  |  |  |  |  |
|  | Unknown vs Normal | 0.803<br>(0.649,0.994) | 0.044 |  |  |
|  | Obese vs Normal | 0.551<br>(0.434,0.701) | 0.000 |  |  |
|  | Overweight vs Normal | 0.678<br>(0.539,0.853) | 0.001 |  |  |
|  | Underweight vs Normal | 0.944<br>(0.523,1.703) | 0.849 |  |  |
| **Comorbidities** |  |  |  |  |  |
|  | Atrial Fibrillation | 1.404<br>(1.047,1.884) | 0.024 |  |  |
|  | Asthma | 0.860<br>(0.682,1.085) | 0.203 |  |  |
|  | Coronary Artery Disease | 1.119<br>(0.891,1.407) | 0.334 |  |  |
|  | Chronic Obstructive Pulmonary Disease | 1.270<br>(1.058,1.524) | 0.010 |  |  |
|  | Depression | 1.006<br>(0.833,1.215) | 0.949 |  |  |
|  | Generalized Anxiety Disorder | 0.772<br>(0.564,1.058) | 0.108 |  |  |
|  | Hepatitis | 0.996<br>(0.807,1.231) | 0.974 |  |  |
|  | Hypertension | 1.061<br>(0.877,1.284) | 0.541 |  |  |
|  | Hyperlipidemia | 0.758<br>(0.630,0.911) | 0.003 |  |  |
|  | Myocardial Infarction | 1.167<br>(0.879,1.550) | 0.286 |  |  |
|  | Peripheral Vascular Disease | 1.246<br>(0.995,1.559) | 0.055 |  |  |
|  | Renal | 1.566<br>(1.237,1.984) | 0.000 |  |  |
|  | Stroke | 1.320<br>(0.991,1.757) | 0.057 |  |  |
|  | Traumatic Brain Injury | 1.059<br>(0.797,1.408) | 0.692 |  |  |
|  | Type 2 Diabetes | 1.196<br>(0.989,1.447) | 0.065 |  |  |
| **Med History** |  |  |  |  |  |
|  | Gabapentin | 1.177<br>(0.958,1.445) | 0.120 |  |  |
|  | Acetaminophen and Codeine | 0.760<br>(0.558,1.035) | 0.081 |  |  |
|  | Acetaminophen and Oxycodone | 0.847<br>(0.677,1.060) | 0.148 |  |  |
|  | Citalopram | 1.176<br>(0.874,1.584) | 0.284 |  |  |
|  | Cyclobenzaprine | 0.949<br>(0.752,1.198) | 0.660 |  |  |
|  | Diazepam | 1.082<br>(0.826,1.416) | 0.568 |  |  |
|  | Diclofenac | 0.892<br>(0.639,1.245) | 0.500 |  |  |
|  | Diphenhydramine | 1.107<br>(0.869,1.410) | 0.412 |  |  |
|  | Enoxaparin | 0.998<br>(0.782,1.274) | 0.988 |  |  |
|  | Hydrochlorothiazide | 0.852<br>(0.681,1.067) | 0.163 |  |  |
|  | Hydromorphone | 0.860<br>(0.672,1.102) | 0.233 |  |  |
|  | Hydroxyzine | 1.280<br>(0.950,1.724) | 0.105 |  |  |
|  | Lorazepam | 1.140<br>(0.916,1.418) | 0.240 |  |  |
|  | Meloxicam | 0.603<br>(0.392,0.927) | 0.021 |  |  |
|  | Metronidazole | 0.900<br>(0.680,1.191) | 0.462 |  |  |
|  | Midazolam | 1.093<br>(0.830,1.441) | 0.526 |  |  |
|  | Naproxen | 0.704<br>(0.547,0.907) | 0.007 |  |  |
|  | Omeprazole | 1.049<br>(0.855,1.287) | 0.647 |  |  |
|  | Ondansetron | 1.272<br>(1.018,1.590) | 0.034 |  |  |
|  | Oxycodone | 1.370<br>(1.093,1.717) | 0.006 |  |  |
|  | Sertraline | 0.930<br>(0.668,1.293) | 0.665 |  |  |
|  | Tramadol | 1.016<br>(0.817,1.264) | 0.885 |  |  |
|  | Trazodone | 1.005<br>(0.781,1.293) | 0.971 |  |  |
|  | Zolpidem | 0.788<br>(0.582,1.066) | 0.122 |  |  |

Prior to PSM, varenicline use was associated with lower hazard of the composite outcome compared with NRT (HR = 0.660, 95% CI: 0.524–0.832, p < 0.001). The association remained statistically significant after PSM (HR = 0.657, 95% CI: 0.501–0.861, p = 0.002), which demonstrated consistency of the association after adjustment for measured confounding.

Covariate associations were generally consistent with those observed of ADRD alone. Older age at index was associated with higher risk (HR = 1.061, 95% CI: 1.048–1.075, p < 0.001). Female sex was associated with lower hazard compared to male sex (HR = 0.825, 95% CI: 0.698–0.976, p = 0.025). Black or African American individuals had a higher hazard compared with White individuals (HR = 1.511, 95% CI: 1.254–1.820, p < 0.001), and Hispanic or Latino ethnicity was associated with higher hazard (HR = 1.864, 95% CI: 1.077–3.227, p = 0.026).

BMI also showed a similar inverse pattern, with both overweight (HR = 0.678, 95% CI: 0.539– 0.853, p = 0.001) and obese (HR = 0.551, 95% CI: 0.434–0.701, p < 0.001) categories associated with lower hazard compared with normal BMI.

Among comorbidities, atrial fibrillation, COPD, and renal disease were associated with higher hazard and statistically significant, while hyperlipidemia was associated with lower hazard.

Prior medication exposures showed mixed association, with some medications (e.g. meloxicam and naproxen) associated with lower hazard and statistically significant, and others (e.g. ondansetron and oxycodone) associated with higher hazard.

### 3.6 Kaplan–Meier Curves

Kaplan–Meier curves also supported the results of the statistical analysis (**Figure 2**, **Figure 3**). For incident ADRD (**Figure 2**), survival curves separated early and grew consistently apart over time, as curves showed separation over time, with higher event-free survival in varenicline users. Similar findings were observed for the composite outcome (**Figure 3**), where varenicline users had a higher probability of being event-free over time.

**Figure 2.**
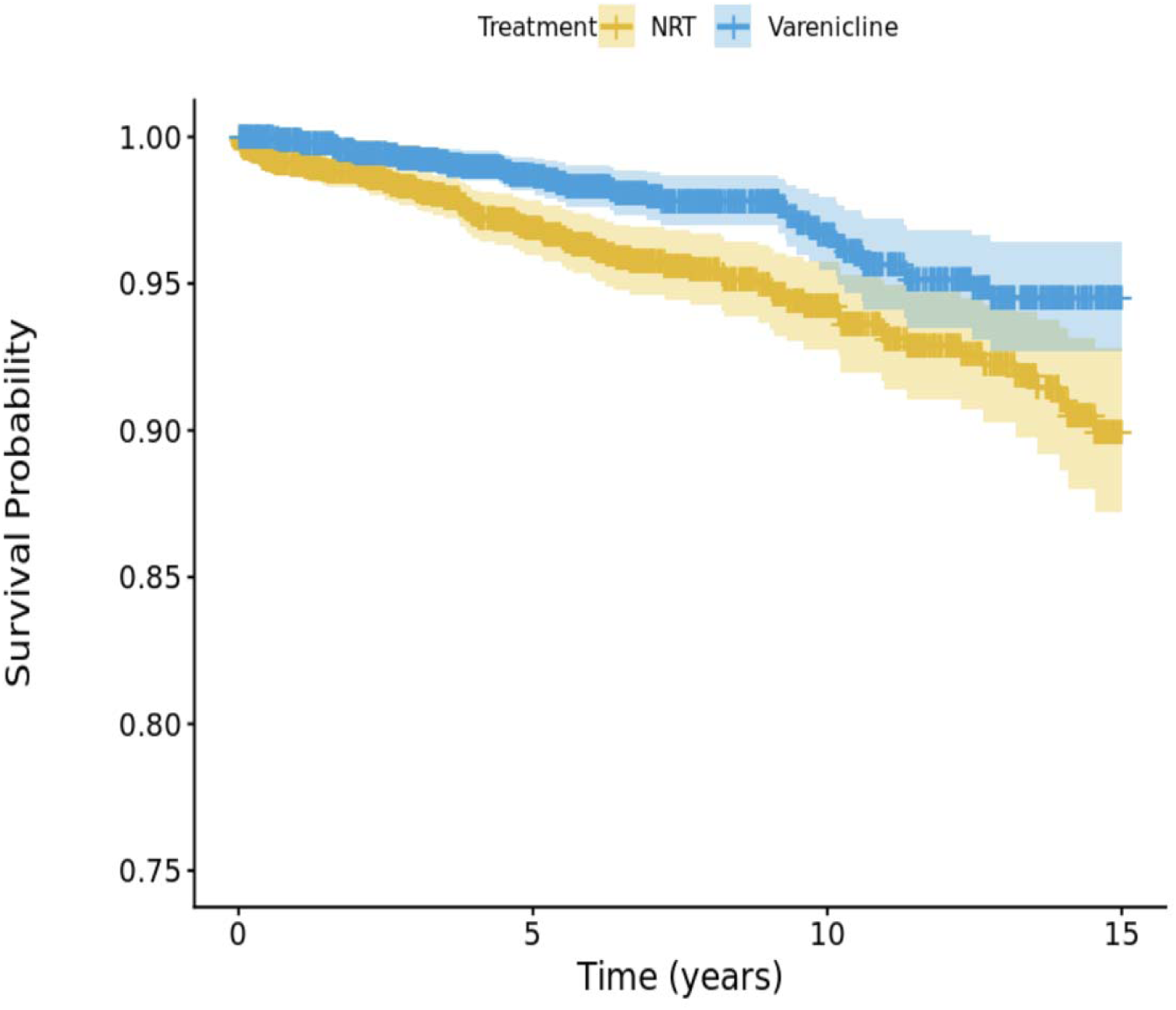
Survival Curves Comparing Varenicline vs. NRT in the Matched Cohort (ADRD)

**Figure 3.**
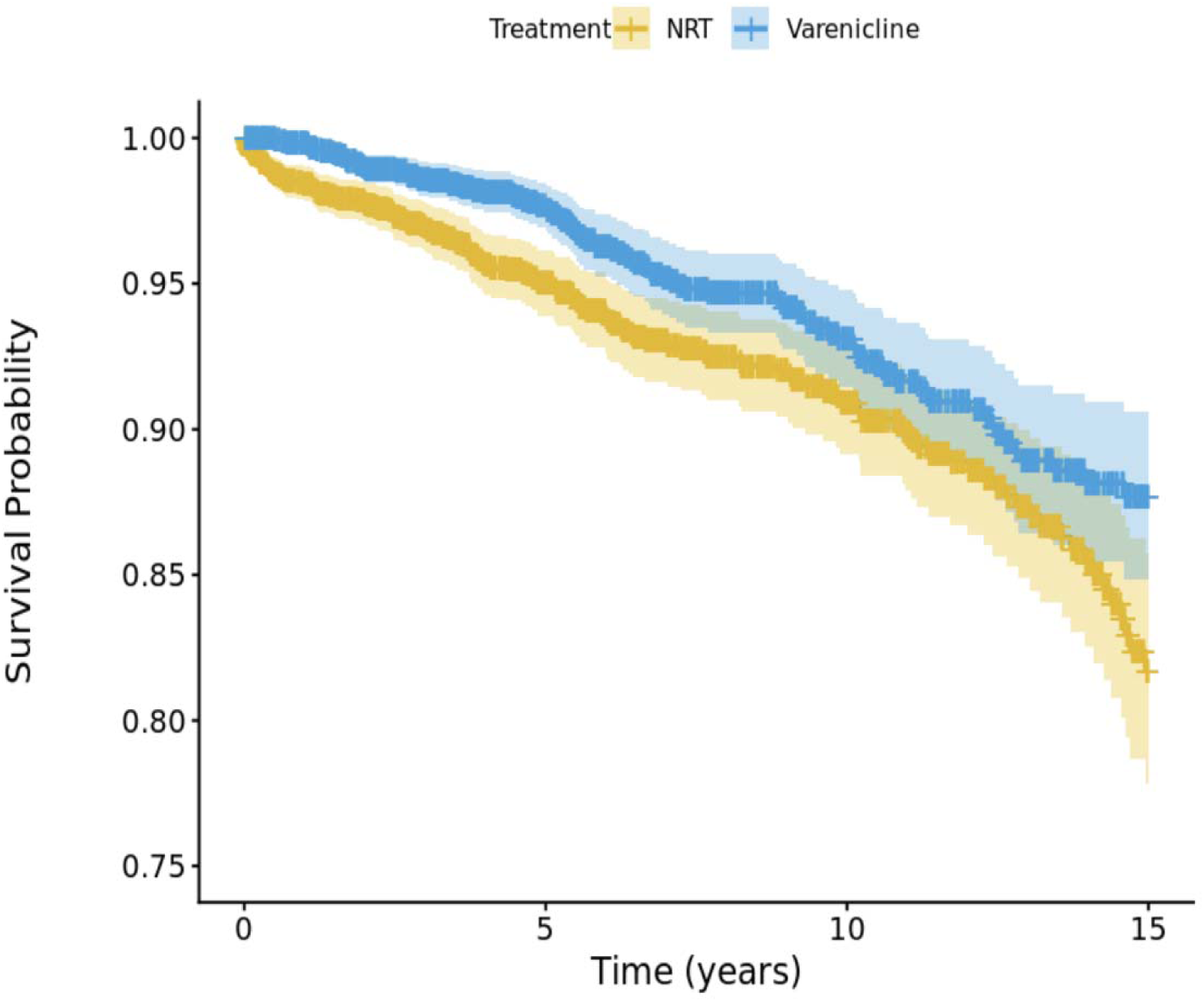
Survival Curves Comparing Varenicline vs. NRT in the Matched Cohort (ADRD or Death)

### 3.7 Number of Prescriptions

In the propensity score–matched cohort (N = 3,516), the mean number of prescriptions was similar between the treatment groups: 3.01 (SD 3.81) for NRT and 2.91 (SD 3.17) for varenicline (**eTable 2A**). The median number of prescriptions was 2 in both groups (interquartile range [IQR]: 1–3). The distribution was right-skewed, with a subset of individuals receiving substantially higher numbers of prescriptions (maximum: 54 for NRT, 28 for varenicline).

The distribution of prescription episodes showed broadly similar patterns between groups (**eTable 2B**). Nearly half of the individuals using NRT (49.15%) and 43.29% of varenicline users had only a single prescription. A higher proportion of varenicline users had 2 to 3 prescriptions (33.11% vs. 26.85% for NRT). The number of individuals with more than three prescriptions was comparable between groups (24.00% for NRT vs. 23.61% for varenicline).

### 3.8 Cumulative Drug Exposure

Cumulative drug exposure, measured in weeks, was higher among varenicline users than among NRT users (**eTable 3A**). Mean exposure duration was 53.88 weeks (SD 74.42) for varenicline and 36.20 weeks (SD 67.26) for NRT. Median exposure duration was also greater in the varenicline group (23.00 weeks; IQR: 8.57–66.00) than in the NRT group (8.57 weeks; IQR: 2.00–35.64), indicating longer treatment duration among varenicline users (**eTable 3B**).

### 3.9 Smoking Status After Treatment

Using condition-based definitions, the proportion of individuals without smoking-related diagnoses was higher among varenicline users than among NRT users (31.40% vs. 25.82%) (**eTable 4A**). Among individuals with smoking-related diagnoses after treatment initiation, the mean time to first smoking-related diagnosis was higher among varenicline users than among NRT users (70.85 vs. 57.11 weeks). Similarly, the mean time to the last smoking-related diagnosis was slightly longer among varenicline users (287.31 vs. 276.39 weeks) (**eTable 4B**).

## 4. Discussion

### 4.1 Principal Findings

This study validated a drug candidate identified through a mechanistically agnostic, explainable AI approach. Prior studies have explored cholinergic pathways.^25–30^ While varenicline has a potential role in dementia, it has received much less attention compared to other candidates such as statins. Our MWAS+ findings position varenicline highly among potential protective agents, supported by observed hazard ratios.

### 4.2 Potential Mechanisms: Smoking Cessation vs Direct Effects

Varenicline’s efficacy in smoking cessation is well established and smoking cessation itself is associated with reduced ADRD risk.^8,31^ A central concern is the extent to which any observed association between varenicline use and reduced risk of ADRD reflects indirect effects mediated through smoking cessation versus a direct pharmacologic benefit of the drug. Smoking cessation is an established modifiable factor associated with reduced dementia risk, and prior large-scale evidence has demonstrated increased ADRD risk among persistent smokers, particularly with greater cumulative exposure.^32,33^ Given that varenicline has consistently been shown to be more effective than NRT in achieving sustained abstinence, any observed protective association may plausibly be driven by its superior cessation efficacy rather than an independent neuroprotective mechanism.^34,35^

### 4.3 Evidence Supporting Superior Smoking Cessation Efficacy of Varenicline

Our results also could be understood in relation to the existing literature indicating that varenicline is generally more effective than NRT for smoking cessation.^36^ Several randomized clinical trials have reported higher abstinence rates in persons with varenicline therapy versus NRT.^13^ In a recent large-scale randomized open-label trial, varenicline resulted in significantly higher rates of continuous abstinence at the end of treatment than transdermal nicotine patch therapy (55.9% vs 43.2%), and participants receiving varenicline also reported less craving, decreased withdrawal symptoms, and decreased smoking satisfaction.^37^ In another recent randomized clinical trial, varenicline sampling effects were reported to lead to greater 6-month self-reported abstinence rates than NRT sampling (17% vs 8%) among adults who smoked and greater reductions in cigarette consumption.^38^ In addition, studies among smokers who initially refused or were not able to quit showed that long-term abstinence rates were significantly increased by varenicline through gradual smoking reduction strategies.^39^ Evidence from sequential treatment trials suggested that switching to varenicline from combined NRT or increasing the dosage of varenicline after a failed initial varenicline therapy might lead to better abstinence outcomes.^40^ Taken together, this evidence suggests that the greater association found with varenicline in our study may at least partly reflect the improved smoking cessation efficacy of varenicline relative to NRT. Because smoking is a well-recognized risk factor of cognitive decline, as well as of ADRD, greater and sustained smoking cessation with varenicline may have contributed to the reduced ADRD risk found in our analyses.

### 4.4 Residual Confounding and Smoking Measurement Limitations

Smoking behavior after treatment could not be comprehensively or systematically ascertained in this study, representing an important source of potential residual confounding. To partially address this, we conducted additional descriptive analyses of prescribing patterns and post-treatment smoking status; however, these measures were limited. While the number of prescriptions was similar between groups, varenicline users exhibited longer cumulative exposure, suggesting greater treatment persistence or adherence. Furthermore, descriptive assessments of post-treatment smoking status showed limited but directionally consistent signals, with a higher proportion of non-smoking status and longer time to smoking-related diagnoses among varenicline users. However, these measures were not systematically captured and should be interpreted cautiously, as they likely represent incomplete and potentially biased proxies of true smoking behavior. Together, these findings offer supportive but not definitive evidence that differences in smoking cessation patterns may contribute to the observed associations.

### 4.5 Possibility of a Direct Neuroprotective Effect

Nevertheless, the magnitude of the hazard ratio observed in our analysis appears to exceed that typically attributable to smoking cessation alone, raising the possibility of an additional direct effect of varenicline on ADRD risk. This interpretation is complicated by the fact that varenicline is frequently used in conjunction with behavioral or supportive interventions, which may further improve cessation outcomes and may introduce additional confounding. Disentangling these pathways is therefore essential, requiring analytic approaches capable of isolating the attributable risk reduction due to smoking cessation from any independent pharmacologic effect of varenicline on ADRD outcomes.^32–35^

### 4.6 Comparison with Prior Evidence

Our findings differ from a prior clinical study conducted in patients with mild-to-moderate Alzheimer’s Disease, which may be explained by short duration, small sample size, and differences in disease stage.^18^ Participants in our study did not have established ADRD at baseline, whereas the Phase II clinical trial conducted by Kim et al. primarily enrolled individuals with more advanced neurodegeneration.^18,41,42^ Emerging preclinical and translational evidence suggests that nicotinic receptor modulation may be more effective during the preclinical or early stages of the disease, when neuronal integrity and synaptic function are relatively preserved, rather than in later stages characterized by substantial neurodegeneration and synaptic loss.^43^ These observations support the hypothesis that nicotinic receptor targeted therapies have greater potential for prevention or early intervention rather than for the treatment of established Alzheimer’s disease.^44,45^

### 4.7 Strengths and Methodological Considerations

To this end, we have included a very large set co-variates (∼400) in the MWAS phase with temporal information and included ∼40 covariates in this cohort analysis. We have employed a new user design with active control and PSM to enhance the statistical rigor.

### 4.8 Clinical Implications and Future Directions

Overall, our findings suggest that future clinical trials should consider varenicline as a potential ADRD prevention agent. Clinicians may also consider varenicline preferentially among smoking cessation therapies for individuals at elevated risk of dementia.

### 4.9 Limitations

Several limitations should be considered when interpreting these findings. First, the number of varenicline users was relatively small after applying the inclusion criteria, which may affect statistical power. Second, the All of Us cohort has been reported to have lower mortality rates than the general population, potentially limiting generalizability. Third, the absence of free-text clinical notes prevented confirmation of whether individuals successfully achieved smoking cessation, which may confound the observed associations. Fourth, the potential for immortal time bias cannot be fully excluded, as the time between cohort entry and the index date was not uniform across participants. Finally, residual confounding may persist despite the use of PSM.

We are currently planning a complementary study using the VA Clinical Data Repository, which offers larger sample sizes and more complete longitudinal records. However, the VA population is predominantly male, which may limit generalizability. Together, findings from All of Us and VA datasets should provide stronger evidence regarding varenicline’s role in ADRD prevention.

## Supporting information

eTable

## Data Availability

This study used data from the All of Us Research Program’s Registered Tier Dataset v8, available to authorized users on the Researcher Workbench.

## Declaration of generative AI and AI-assisted technologies in the manuscript preparation process

During the preparation of this work the author(s) used ChatGPT in order to improve language and readability. After using this tool/service, the author(s) reviewed and edited the content as needed and take(s) full responsibility for the content of the published article.

## Acknowledgement

We gratefully acknowledge All of Us participants for their contributions, without whom this research would not have been possible. We also thank the National Institutes of Health’s All of Us Research Program for making available the participant data examined in this study.

## Reference

1. Cheng Y, Zamrini E, Ahmed A, Wu W-C, Shao Y, Zeng-Treitler Q. Medication-Wide Association Study Plus (MWAS+): A Proof of Concept Study on Drug Repurposing. Medical Sciences. 08/31 2022;10:48. doi:10.3390/medsci10030048

2. Zeng Q, Shao Y, Yin Y, Zamrini E, Ahmed A, Yan C. Hybrid Value-Aware Transformer Identifies Novel and Suspected Drugs for Alzheimer’s Disease. Innovation in Aging. 2025;9(Supplement_2)doi:10.1093/geroni/igaf122.3958

3. Shao Y, Cheng Y, Nelson SJ, et al. Hybrid Value-Aware Transformer Architecture for Joint Learning from Longitudinal and Non-Longitudinal Clinical Data. J Pers Med. Jun 29 2023;13(7)doi:10.3390/jpm13071070

4. Shao Y, Cheng Y, Shah RU, Weir CR, Bray BE, Zeng-Treitler Q. Shedding Light on the Black Box: Explaining Deep Neural Network Prediction of Clinical Outcomes. J Med Syst. Jan 4 2021;45(1):5. doi:10.1007/s10916-020-01701-8

5. Ali Ahmed BD, Prakash Deedwania, Daniel Taub, Andrew Zullo, Charles Faselis, Jose Vargas, Yijun Shao, Stuart Nelson, Qing Zeng-Treitler. 5089.0 - A novel framework to evaluate drugs for repurposing against dementia. presented at: American Public Health Association 2025 Annual Meeting; November 5 2025; Washington, D.C. Session Chronic Disease Management and Stigma in Public Health. https://apha.confex.com/apha/2025/meetingapp.cgi/Paper/583056

6. Hampel H, Mesulam MM, Cuello AC, et al. The cholinergic system in the pathophysiology and treatment of Alzheimer’s disease. Brain. Jul 1 2018;141(7):1917–1933. doi:10.1093/brain/awy132

7. Carlson AB, Kraus GP. Physiology, Cholinergic Receptors. StatPearls. StatPearls Publishing Copyright © 2026, StatPearls Publishing LLC.; 2026.

8. Durazzo TC, Mattsson N, Weiner MW. Smoking and increased Alzheimer’s disease risk: a review of potential mechanisms. Alzheimers Dement. Jun 2014;10(3 Suppl):S122–45. doi:10.1016/j.jalz.2014.04.009

9. Krivanek TJ, Gale SA, McFeeley BM, Nicastri CM, Daffner KR. Promoting Successful Cognitive Aging: A Ten-Year Update. J Alzheimers Dis. 2021;81(3):871–920. doi:10.3233/jad-201462

10. Hersi M, Beck A, Hamel C, et al. Effectiveness of smoking cessation interventions among adults: an overview of systematic reviews. Syst Rev. Jul 12 2024;13(1):179. doi:10.1186/s13643-024-02570-9

11. Kelley DE, Boynton MH, Noar SM, et al. Effective Message Elements for Disclosures About Chemicals in Cigarette Smoke. Nicotine Tob Res. Aug 14 2018;20(9):1047–1054. doi:10.1093/ntr/ntx109

12. Tonstad S, Arons C, Rollema H, et al. Varenicline: mode of action, efficacy, safety and accumulated experience salient for clinical populations. Curr Med Res Opin. May 2020;36(5):713–730. doi:10.1080/03007995.2020.1729708

13. Altalhi I. Effectiveness of varenicline versus nicotine replacement therapy in smoking cessation: A systematic review. journal article. Tobacco Prevention & Cessation. 2024;10(Supplement 1)doi:10.18332/tpc/194412

14. Seyedaghamiri F, Hosseini L, Kazmi S, et al. Varenicline improves cognitive impairment in a mouse model of mPFC ischemia: The possible roles of inflammation, apoptosis, and synaptic factors. Brain Res Bull. Apr 2022;181:36–45. doi:10.1016/j.brainresbull.2022.01.010

15. Lilja AM, Porras O, Storelli E, Nordberg A, Marutle A. Functional interactions of fibrillar and oligomeric amyloid-β with alpha7 nicotinic receptors in Alzheimer’s disease. J Alzheimers Dis. 2011;23(2):335–47. doi:10.3233/jad-2010-101242

16. Athari SZ, Kazmi S, Vatandoust SM, et al. Varenicline Attenuates Memory Impairment in Amyloid-Beta-Induced Rat Model of Alzheimer’s Disease. Neurochem Res. Jan 27 2025;50(2):86. doi:10.1007/s11064-025-04338-6

17. Ni R, Marutle A, Nordberg A. Modulation of α7 nicotinic acetylcholine receptor and fibrillar amyloid-β interactions in Alzheimer’s disease brain. J Alzheimers Dis. 2013;33(3):841–51. doi:10.3233/jad-2012-121447

18. Kim SY, Choi SH, Rollema H, et al. Phase II crossover trial of varenicline in mild-to-moderate Alzheimer’s disease. Dement Geriatr Cogn Disord. 2014;37(3-4):232–45. doi:10.1159/000355373

19. All of Us Research Program. https://allofus.nih.gov/

20. Dugger BN, Dickson DW. Pathology of Neurodegenerative Diseases. Cold Spring Harb Perspect Biol. Jul 5 2017;9(7)doi:10.1101/cshperspect.a028035

21. Merritt VC, Zhang R, Sherva R, et al. Curation and validation of electronic medical record-based dementia diagnoses in the VA Million Veteran Program. J Alzheimers Dis. Jan 2025;103(1):180–193. doi:10.1177/13872877241299130

22. Cheng Y, Ahmed A, Zamrini E, Tsuang DW, Sheriff HM, Zeng-Treitler Q. Alzheimer’s Disease and Alzheimer’s Disease-Related Dementias in Older African American and White Veterans. J Alzheimers Dis. 2020;75(1):311–320. doi:10.3233/jad-191188

23. Austin PC. Optimal caliper widths for propensity-score matching when estimating differences in means and differences in proportions in observational studies. Pharm Stat. Mar-Apr 2011;10(2):150–61. doi:10.1002/pst.433

24. Austin PC. Using the Standardized Difference to Compare the Prevalence of a Binary Variable Between Two Groups in Observational Research. Communications in Statistics - Simulation and Computation. 2009/05/14 2009;38(6):1228–1234. doi:10.1080/03610910902859574

25. Gray SL, Anderson ML, Dublin S, et al. Cumulative use of strong anticholinergics and incident dementia: a prospective cohort study. JAMA Intern Med. Mar 2015;175(3):401–7. doi:10.1001/jamainternmed.2014.7663

26. Gray SL, Anderson ML, Hanlon JT, et al. Exposure to Strong Anticholinergic Medications and Dementia-Related Neuropathology in a Community-Based Autopsy Cohort. J Alzheimers Dis. 2018;65(2):607–616. doi:10.3233/jad-171174

27. Gray SL, Hanlon JT. Anticholinergic medication use and dementia: latest evidence and clinical implications. Ther Adv Drug Saf. Oct 2016;7(5):217–224. doi:10.1177/2042098616658399

28. Gray SL, Walker R, Dublin S, et al. Histamine-2 receptor antagonist use and incident dementia in an older cohort. J Am Geriatr Soc. Feb 2011;59(2):251–7. doi:10.1111/j.1532-5415.2010.03275.x

29. Gray SL, Walker RL, Dublin S, et al. Proton Pump Inhibitor Use and Dementia Risk: Prospective Population-Based Study. J Am Geriatr Soc. Feb 2018;66(2):247–253. doi:10.1111/jgs.15073

30. Marcum ZA, Hohl SD, Barthold D, Zaslavsky O, Larson EB, Gray SL. Beliefs about benefits and harms of medications and supplements for brain health. Prev Med Rep. Mar 2020;17:101060. doi:10.1016/j.pmedr.2020.101060

31. Burke MV, Hays JT, Ebbert JO. Varenicline for smoking cessation: a narrative review of efficacy, adverse effects, use in at-risk populations, and adherence. Patient Prefer Adherence. 2016;10:435–41. doi:10.2147/ppa.S83469

32. Livingston G, Huntley J, Sommerlad A, et al. Dementia prevention, intervention, and care: 2020 report of the Lancet Commission. Lancet. Aug 8 2020;396(10248):413–446. doi:10.1016/s0140-6736(20)30367-6

33. Rusanen M, Kivipelto M, Quesenberry CP, Jr., Zhou J, Whitmer RA. Heavy smoking in midlife and long-term risk of Alzheimer disease and vascular dementia. Arch Intern Med. Feb 28 2011;171(4):333–9. doi:10.1001/archinternmed.2010.393

34. Cahill K, Stevens S, Perera R, Lancaster T. Pharmacological interventions for smoking cessation: an overview and network meta-analysis. Cochrane Database Syst Rev. May 31 2013;2013(5):Cd009329. doi:10.1002/14651858.CD009329.pub2

35. Anthenelli RM, Benowitz NL, West R, et al. Neuropsychiatric safety and efficacy of varenicline, bupropion, and nicotine patch in smokers with and without psychiatric disorders (EAGLES): a double-blind, randomised, placebo-controlled clinical trial. Lancet. Jun 18 2016;387(10037):2507–20. doi:10.1016/s0140-6736(16)30272-0

36. Taylor GMJ, Taylor AE, Thomas KH, et al. The effectiveness of varenicline versus nicotine replacement therapy on long-term smoking cessation in primary care: a prospective cohort study of electronic medical records. Int J Epidemiol. Dec 1 2017;46(6):1948–1957. doi:10.1093/ije/dyx109

37. Aubin H-J, Bobak A, Britton JR, et al. Varenicline versus transdermal nicotine patch for smoking cessation: results from a randomised open-label trial. Thorax. 2008;63(8):717–724. doi:10.1136/thx.2007.090647

38. Carpenter MJ, Smith TT, Wahlquist AE, et al. Medication Samples and Smoking Cessation Among Adults: A Randomized Clinical Trial. JAMA Netw Open. May 1 2026;9(5):e2611418. doi:10.1001/jamanetworkopen.2026.11418

39. Ebbert JO, Hughes JR, West RJ, et al. Effect of varenicline on smoking cessation through smoking reduction: a randomized clinical trial. Jama. Feb 17 2015;313(7):687–94. doi:10.1001/jama.2015.280

40. Cinciripini PM, Green CE, Shete S, et al. Smoking Cessation After Initial Treatment Failure With Varenicline or Nicotine Replacement: A Randomized Clinical Trial. Jama. May 28 2024;331(20):1722–1731. doi:10.1001/jama.2024.4183

41. Drummond E, Wisniewski T. Alzheimer’s disease: experimental models and reality. Acta Neuropathol. Feb 2017;133(2):155–175. doi:10.1007/s00401-016-1662-x

42. Polis B, Samson AO. Addressing the Discrepancies Between Animal Models and Human Alzheimer’s Disease Pathology: Implications for Translational Research. J Alzheimers Dis. 2024;98(4):1199–1218. doi:10.3233/jad-240058

43. Terry AV, Jr., Buccafusco JJ. The cholinergic hypothesis of age and Alzheimer’s disease-related cognitive deficits: recent challenges and their implications for novel drug development. J Pharmacol Exp Ther. Sep 2003;306(3):821–7. doi:10.1124/jpet.102.041616

44. Parri HR, Hernandez CM, Dineley KT. Research update: Alpha7 nicotinic acetylcholine receptor mechanisms in Alzheimer’s disease. Biochem Pharmacol. Oct 15 2011;82(8):931–42. doi:10.1016/j.bcp.2011.06.039

45. Buckingham SD, Jones AK, Brown LA, Sattelle DB. Nicotinic acetylcholine receptor signalling: roles in Alzheimer’s disease and amyloid neuroprotection. Pharmacol Rev. Mar 2009;61(1):39–61. doi:10.1124/pr.108.000562

