## Supplementary material for "Varenicline as a Repurposable Drug Candidate for ADRD Prevention": eTable

| **eTable 1. ICD Diagnosis Codes** | | | |
| --- | --- | --- | --- |
|  | **ICD-9-CM** | **ICD-10** | **Enhanced ICD-9-CM** |
| Alzheimer's Disease and Related Dementias | 290.0, 290.10, 290.40, 290.41, 290.42, 290.43, 291.2, 292.82, 294.10, 294.11, 294.11/042, 294.11/331.5, 294.11/332.0, 294.11/333.4, 294.20, 294.20/042, 294.20/331.5, 294.20/332.0, 294.20/333.4, 294.21, 294.8, 331.0, 331.19, 331.2  331.7, 331.82, 331.89, 331.9 | F01.50, F01.51, F02.80, F02.80/B20, F02.80/G10, F02.80/G20, F02.80/G91.2, F02.81, F02.81/B20, F02.81/G10, F02.81/G20, F02.81/G91.2, F03.90, F03.91, F10.27, F10.97, F13.27, F13.97, F18.17, F18.27, F18.97, F19.17, F19.27, F19.97, G30.0, G30.1, G30.8, G30.9, G31.09, G31.83 |  |
| Atrial Fibrillation | 427.31x, | I48.0x, I48.1x, I48.2x, I48.91x |  |
| Asthma | 493.0x, 493.1x, 493.2x, 493.8x, 493.9x | J45.2x, J45.3x, J45.4x, J45.5x, J45.9x |  |
| Coronary Artery Disease | 414 | I25 |  |
| Chronic Obstructive Pulmonary Disease | 491.x, 496.x | J41.x, J42.x, J44.x |  |
| Depression | 300.4, 301.12, 309.0, 309.1, 311 | F20.4, F31.3–F31.5, F32.x, F33.x, F34.1, F41.2, F43.2 | 296.2, 296.3, 296.5, 300.4, 309.x, 311 |
| Generalized Anxiety Disorder | 300.02 | F41.1 |  |
| Hepatitis | 070-070.9, 571.40, 571.41, 571.42, 571.49, 571.1, 573.3, V12.09 | B15.x–B19.x, K73.x, K74.69, Z86.19 |  |
| Hypertension | 401.1, 401.9, 642.0, 401.0, 402.x–405.x, 642.1, 642.2, 642.7, 642.9 | I10.x, I11.x–I13.x, I15.x | 401.x 402.x–405.x |
| Hyperlipidemia | 272.0x, 272.1x, 272.2x, 272.3x, 272.4x | E78.0x, E78.1x, E78.2x, E78.3x, E78.4x, E78.5x |  |
| Myocardial Infarction | 410.x, 412.x | I21.x, I22.x, I25.2 |  |
| Peripheral vascular disease | 443.9, 441.x, 785.4, V43.4, Procedure 38.48 | I70.x, I71.x, I73.1, I73.8, I73.9, I77.1, I79.0, I79.2, K55.1, K55.8, K55.9, Z95.8, Z95.9 | 093.0, 437.3, 440.x, 441.x, 443.1–443.9, 47.1, 557.1, 557.9, V43.4 |
| Renal disease | 582.x, 583–583.7, 585.x, 586.x, 588.x | I12.0, I13.1, N03.2–N03.7, N05.2– N05.7, N18.x, N19.x, N25.0, Z49.0– Z49.2, Z94.0, Z99.2 | 403.01, 403.11, 403.91, 404.02, 404.03, 404.12, 404.13, 404.92, 404.93, 582.x, 583.0–583.7, 585.x, 586.x, 588.0, V42.0, V45.1, V56.x |
| Stroke | 433.01x, 433.11x, 433.21x, 433.31x, 433.81x, 433.91x, 434.x, V12.54x | I63.x, I69.3x, |  |
| Type 2 Diabetes | 250.2, 250.20, 250.22, 250.32, 250.4, 250.40, 250.42, 250.5, 250.50, 250.52, 250.6, 250.60, 250.62, 250.7, 250.70, 250.72, 250.8, 250.80, 250.30, 250.02, 250.10, 250.82, 250.12, 250.9, 250.90, 250.92, 250.0, 250.00 | E11, E11.0, E11.00, E11.01, E11.21, E11.22, E11.29, E11.3, E11.31, E11.311, E11.319, E11.32, E11.321, E11.329, E11.33, E11.331, E11.339, E11.34, E11.341, E11.349, E11.35, E11.351, E11.359, E11.36, E11.39, E11.4, E11.40, E11.41, E11.42, E11.43, E11.44, E11.49, E11.5, E11.51, E11.52, E11.59, E11.6, E11.61, E11.610, E11.618, E11.62, E11.620, E11.621, E11.622, E11.628, E11.63, E11.630, E11.638, E11.64, E11.641, E11.649, E11.65, E11.69, E11.8, E11.9 |  |
| Traumatic Brain Injury | 800.x, 801.x, 803.x, 804.x, 850.x-854.x, 950.1-950.3, 959.01, 310.2, 905.0, 907.0, 959.9, V15.52 | Z87.820, S02.0, S02.10Xx, S02.110-S02.119, S02.19, S02.8Xx, S02.9, S04.02-S04.04, S06.0X0-S06.0X9, S06.1X0-S06.1X9, S06.2X0-S06.2X9, S06.300-S06.309, S06.310-S06.319, S06.320-S06.329, S06.330-S06.339, S06.340-S06.349, S06.350-S06.359, S06.360-S06.369, S06.370-S06.379, S06.380-S06.389, S06.4X0-S06.4X9, S06.5X0-S06.5X9, S06.6X0-S06.6X9, S06.890-S06.899, S06.9X0-S06.9X9, S07.1 |  |

| **eTable 2A. Descriptive Statistics of Prescription Counts by Smoking Cessation Therapy** | | | | | | | | | |
| --- | --- | --- | --- | --- | --- | --- | --- | --- | --- |
|  | **N** | **Mean** | **SD** | **Median** | **Q1** | **Q3** | **Min** | **Max** | **%** |
| **Therapy** |  |  |  |  |  |  |  |  |  |
| NRT | 1758 | 3.01 | 3.81 | 2 | 1 | 3 | 1 | 54 | 50 |
| Varenicline | 1758 | 2.91 | 3.17 | 2 | 1 | 3 | 1 | 28 | 50 |

| **eTable 2B. Distribution of Prescriptions by Smoking Cessation Therapy** | | | |
| --- | --- | --- | --- |
|  | **Episodes** | **N** | **%** |
| **Therapy** |  |  |  |
| NRT | 1 | 864 | 49.15 |
|  | 2 | 301 | 17.12 |
|  | 3 | 171 | 9.73 |
|  | >3 | 422 | 24.00 |
| Varenicline | 1 | 761 | 43.29 |
|  | 2 | 363 | 20.65 |
|  | 3 | 219 | 12.46 |
|  | >3 | 415 | 23.61 |

| **eTable 3A. Descriptive Statistics of Cumulative Drug Exposure (Weeks) by Therapy** | | | | | | | | | |
| --- | --- | --- | --- | --- | --- | --- | --- | --- | --- |
|  | **N** | **Mean** | **SD** | **Median** | **Q1** | **Q3** | **Min** | **Max** | **%** |
| **Therapy** |  |  |  |  |  |  |  |  |  |
| NRT | 1758 | 36.20 | 67.26 | 8.57 | 2.00 | 35.64 | 0 | 535.86 | 50 |
| Varenicline | 1758 | 53.88 | 74.42 | 23.00 | 8.57 | 66.00 | 0 | 452.14 | 50 |

| **eTable 3B. Categorized Distribution of Cumulative Drug Exposure (Weeks) by Therapy Group** | | | | |
| --- | --- | --- | --- | --- |
|  | **Time** | **N** | **Mean Time** | **%** |
| **Therapy** |  |  |  |  |
| NRT | ≥0 and ≤4 | 521 | 1.36 | 29.64 |
|  | >4 and ≤12 | 345 | 6.74 | 19.62 |
|  | >12 and ≤24 | 189 | 16.50 | 10.75 |
|  | >24 | 516 | 98.30 | 29.35 |
|  | NA | 187 | — | 10.64 |
| Varenicline | ≥0 and ≤4 | 198 | 1.41 | 11.26 |
|  | >4 and ≤12 | 272 | 7.47 | 15.47 |
|  | >12 and ≤24 | 305 | 16.55 | 17.35 |
|  | >24 | 739 | 100.43 | 42.04 |
|  | NA | 244 | — | 13.88 |

| **eTable 4A. Time to First Smoking-Related Diagnosis After Treatment by Therapy** | | | | |
| --- | --- | --- | --- | --- |
|  | **Smoking Status** | **N** | **Mean Follow-up First Smoking Diagnosis Duration (weeks)** | **%** |
| **Therapy** |  |  |  |  |
| NRT | No | 454 | — | 25.82 |
|  | Yes | 1304 | 57.11 | 74.18 |
| Varenicline | No | 552 | — | 31.40 |
|  | Yes | 1206 | 70.85 | 68.60 |

| **eTable 4B. Time to Last Smoking-Related Diagnosis After Treatment by Therapy** | | | | |
| --- | --- | --- | --- | --- |
|  | **Smoking Status** | **N** | **Mean Follow-up Last Smoking Diagnosis Duration (weeks)** | **%** |
| **Therapy** |  |  |  |  |
| NRT | No | 454 | — | 25.82 |
|  | Yes | 1304 | 276.39 | 74.18 |
| Varenicline | No | 552 | — | 31.40 |
|  | Yes | 1206 | 287.31 | 68.60 |
